# SARS-CoV-2 lethality did not change over time in two Italian Provinces

**DOI:** 10.1101/2020.05.23.20110882

**Authors:** Maria Elena Flacco, Cecilia Acuti Martellucci, Francesca Bravi, Giustino Parruti, Alfonso Mascitelli, Lorenzo Mantovani, Stefania Boccia, Lamberto Manzoli

**Author notes:** **Corresponding author:** Lamberto Manzoli, University of Ferrara, Vecchi Istituti Biologici - VIB, Via Fossato di Mortara 64B - 44121 Ferrara, Italy. These authors equally contributed to this manuscript.

## Abstract

**Background:** Some experts recently reported that SARS-CoV-2 lethality decreased considerably, but no evidence is yet available. This retrospective cohort study aimed to evaluate whether SARS-CoV-2 case-fatality rate decreased with time, adjusting for several potential confounders.

**Methods:** We included all subjects diagnosed with SARS-CoV-2 infection in Ferrara and Pescara provinces, Italy. Information were collected from local registries, clinical charts, and electronic health records. We compared the case-fatality rate (after ≥28 days of follow-up) of the subjects diagnosed during April and March, 2020. We used Cox proportional hazards analysis and random-effect logistic regression, adjusting for age, gender, hypertension, type II diabetes, major cardiovascular diseases (CVD), chronic obstructive pulmonary diseases (COPD), cancer and renal disease.

**Results:** The sample included 2493 subjects (mean age 58.6y; 47.7% males). 258 persons deceased, after a mean of 16.1 days of follow-up. The mean age of those who died substantially increased from March (78.1±11.0y) to April (84.3±10.2y). From March to April, the case-fatality rate did not decrease in the total sample (9.5% versus 12.1%; adjusted hazard ratio 0.93; 95% Confidence Interval: 0.71-1.21; p=0.6), and in any age-class.

**Conclusions:** In this sample, SARS-CoV-2 case-fatality rate did not decrease over time, in contrast with recent claims of a substantial improvement of SARS-CoV-2 clinical management. The findings require confirmation from larger datasets.

**Author summary:** *Why was this study done?:*

- Some experts recently reported that SARS-CoV-2 lethality decreased considerably, but no evidence is yet available.

*What did the researchers do and find?:*

- We carried out a retrospective cohort study on 2493 SARS-CoV-2 infected subjects from two Italian provinces, evaluating the potential variation of the case-fatality rate over time.
- From March to April, SARS-CoV-2 case-fatality rate did not decrease, overall and in any age-class.

*What do these findings mean?:*

- The therapies and clinical management of SARS-CoV-2 infected subjects did not determine a substantial change of the clinical course of the disease from March to April, 2020.

## Introduction

As of mid-July, 2020, SARS-CoV-2 pandemic has caused more than 500,000 deaths worldwide [1], with largely discrepant case-fatality rates across countries (from <1% to 16%) [2], likely due to differences in population age structure [3, 4], variations in testing policies and case recording [5], and/or preparedness of the healthcare system, which in turn is affected by the intensity of the spread [4, 5].

Since the start of the pandemic, Italy has been among the countries with the highest death toll, with nearly 35,000 recorded deaths [1], and an estimated case-fatality rate of 14% [2], which peaked at 20% among the citizens aged ≥80 years [6].

In the last months, it has been suggested that SARS-CoV-2 lethality may have decreased, mostly as a consequence of more tailored therapeutic approaches [7-11]. Although the claims were made by physicians actively engaged in the care of infected patients, the available evidence is anecdotal or based upon case-studies.

We analysed the data of all infected cases in two Italian Provinces to evaluate whether SARS-CoV-2 case-fatality rate decreased with time, adjusting for main potential confounders.

## Methods

This retrospective cohort study included all subjects with a diagnosed SARS-CoV-2 infection in the Provinces of Ferrara and Pescara, between March 3 (the onset date of the first cases), and May 3, 2020. All participants were followed up to May 31, 2020. All infections were diagnosed by the central laboratories of Ferrara University Hospital or Pescara Hospital through RT-PCR (Reverse transcription polymerase chain reaction) test on nasopharyngeal swabs, and were confirmed by the Italian National Institute of Health. Information on age, gender, and comorbidities of all participants were collected from local registries, clinical charts (for hospitalized patients), and through data-linkage with hospital discharge abstracts (Italian SDO) and the National database of drug prescription. Electronic databases were queried from the day of the diagnosis until January 1st, 2015. All data have been revised manually by two physicians (LM and MEF), and the following conditions have been included in the analyses: hypertension, type II diabetes, major cardiovascular diseases (heart failure, myocardial infarction and stroke - CVD), chronic obstructive pulmonary diseases (COPD, bronchitis, pneumonia, asthma, and emphysema), malignant tumors and renal disease. No additional data were requested or collected.

The study complies with the Declaration of Helsinki, the research protocol was approved by the Ethics Committee of the Emilia-Romagna Region (code 287, approved on March 24, 2020), and the requirement for informed consent was waived because of the retrospective and pseudo-anonymized nature of the data.

### Data Analysis

We compared the case-fatality rate (fatal / confirmed cases) during the first 29 days after the index day (from March 3 to March 31) with that of the second half of the period (days 30-63; from April 1 to May 3). For the sake of simplicity, the second period, composed by April and a few days of May, will be defined as “April” from now on. The differences between the two periods were initially evaluated using t-test for continuous variables, and chi-squared test for categorical ones. The potential independent predictors of death were then evaluated using Cox proportional hazards analysis (censoring at May 31, to include ≥28 days of follow-up). All covariates were included a priori in the model in their original form, with the exception of age, which was treated as either continuous or ordinal, to explore the association between the outcome and several age classes. Schoenfeld’s test was used to assess the validity of proportional hazards assumption, and Nelson-Aalen cumulative hazard estimates to check the validity of constant incidence ratios during the follow-up [12]. Using the data censored at 28 days of follow-up, a random-effect logistic regression was also fit, with province as the cluster unit. The same above criteria were used to build the final model. Missing data were <5% in all primary analyses; therefore, no missing imputation technique was adopted. Statistical significance was defined as a two-sided p-value<0.05, and all analyses were carried out using Stata, version 13.1 (Stata Corp., College Station, TX, 2014). The same analytical approach was adopted to assess a previous dataset [13], which was based upon the samples collected up to April 25, using a shorter, 10-days follow-up period. The present analyses have been updated with the new data and a longer, 28-days follow-up.

## Results

The sample consisted of 2493 subjects (mean age 58.6y; 47.7% males); of them, 33.0% were hypertensive, 13.0% diabetics, and 15.5% with CVD. Some of the characteristics of the sample significantly varied from March to April. Infected subjects were older by 3.1 years, and the proportion of females, diabetics, subjects with CVD, cancer and renal diseases significantly increased (Table 1).

**Table 1.** Characteristics of the sample, overall and by time of SARS-CoV-2 infection diagnosis after the first case (March 3, 2020).

|  | **Total**<br>**sample**<br>**(n=2493)** | **March**<br>**2020<sup>a</sup>**<br>**(n=1658)** | **April 1-May 3**<br>**2020<sup>b</sup>**<br>**(n=835)** | **p\*** |
| --- | --- | --- | --- | --- |
| Mean age (SD), years | 58.6 (21.1) | 57.6 (19.5) | 60.7 (24.0) | <0.001 |
| Age-class in years, % |  |  |  |  |
| - <18 | 3.6 | 3.1 | 4.4 | 0.10 |
| - 18-39.9 | 15.1 | 14.2 | 16.9 | 0.074 |
| - 40-49.9 | 14.5 | 16.1 | 11.3 | 0.001 |
| - 50-59.9 | 17.6 | 18.8 | 15.3 | 0.034 |
| - 60-69.9 | 16.1 | 19.0 | 10.4 | <0.001 |
| - 70-79.9 | 13.2 | 14.2 | 11.1 | 0.031 |
| - ≥80 | 19.9 | 14.6 | 30.5 | <0.001 |
| Male gender, % | 47.7 | 51.8 | 39.5 | <0.001 |
| Hypertension, % | 33.0 | 33.5 | 32.1 | 0.5 |
| Diabetes, % | 13.0 | 11.3 | 16.2 | 0.001 |
| Major cardiovascular diseases, % | 15.5 | 14.1 | 18.4 | 0.004 |
| COPD, % | 5.1 | 5.4 | 4.7 | 0.5 |
| Cancer, % | 7.1 | 8.0 | 5.4 | 0.016 |
| Renal diseases, % | 5.3 | 4.3 | 7.4 | 0.001 |

<sup>a</sup> From March 3 to March 31, 2020.
<sup>b</sup> From April 1 to May 3, 2020.
\* Chi-squared test for categorical variables, t-test for continuous ones.

Overall, 258 persons deceased (after a mean of 16.1 days of follow-up): 157 of the 1658 subjects diagnosed in March (9.5%), 101 of the 835 subjects diagnosed from April 1 to May 3 (12.1%). The mean age of those who died substantially increased: it was 78.1±11.0 for those diagnosed in March, 84.3±10.2 for those detected in April (p<0.001). In March, 33 of those deceased were younger than 70 years, and 10 were younger than 60 years. In April, six deaths occurred in subjects younger than 70 years (two among those younger than 60y).

As shown in Table 2, in the overall sample the crude SARS-CoV-2 lethality significantly increased from March to April (from 9.5% to 12.1%; p=0.042). The increase was significant among the males (from 9.9% to 15.8%; p=0.005), and those without hypertension (from 4.4% to 6.7%; p=0.040), diabetes (from 7.4% to 10.7%; p=0.005), COPD (from 8.5% to 11.1%; p=0.040) and cancer (from 8.7% to 11.3%; p=0.050). No significant change were observed among the other categories of subjects.

**Table 2.** Proportion of deaths, overall and by time of SARS-CoV-2 infection diagnosis after the first case (March 3, 2020), and hazard ratios (HRs) predicting the time to death of patients diagnosed in April vs March 2020.

|  | Total sample | March 2020 <sup>a</sup> | April 1-May 3 2020 <sup>b</sup> | p <sup>c</sup> | April vs March HR (95% CI) | p <sup>d</sup> |
| --- | --- | --- | --- | --- | --- | --- |
| Overall | 10.3 | 9.5 | 12.1 | 0.042 | 0.93 (0.71-1.21) | 0.6 |
| Age-class in years, % |  |  |  |  |  |  |
| - <18 | 0.0 | 0.0 | 0.0 | -- | -- | -- |
| - 18-39.9 | 0.3 | 0.0 | 0.7 | 0.2 | -- | -- |
| - 40-49.9 | 0.6 | 0.8 | 0.0 | 0.4 | -- | -- |
| - 50-59.9 | 2.1 | 2.6 | 0.8 | 0.2 | 0.22 (0.02-2.05) | 0.2 |
| - 60-69.9 | 6.7 | 7.3 | 4.6 | 0.4 | 0.66 (0.22-1.99) | 0.5 |
| - 70-79.9 | 18.8 | 19.1 | 18.3 | 0.9 | 0.92 (0.52-1.63) | 0.8 |
| - ≥80 | 31.6 | 32.6 | 30.6 | 0.6 | 0.94 (0.67-1.30) | 0.7 |
| Gender, % |  |  |  |  |  |  |
| - Females | 9.3 | 9.0 | 9.7 | 0.7 | 0.65 (0.44-0.95) | 0.027 |
| - Males | 11.5 | 9.9 | 15.8 | 0.005 | 1.12 (0.77-1.61) | 0.6 |
| Hypertension, % |  |  |  |  |  |  |
| - No | 5.2 | 4.4 | 6.7 | 0.040 | 0.92 (0.58-1.45) | 0.7 |
| - Yes | 20.9 | 19.6 | 23.5 | 0.2 | 0.85 (0.61-1.19) | 0.3 |
| Diabetes, % |  |  |  |  |  |  |
| - No | 8.3 | 7.4 | 10.7 | 0.005 | 1.03 (0.75-1.42) | 0.8 |
| - Yes | 24.2 | 27.7 | 19.3 | 0.08 | 0.57 (0.34-0.94) | 0.028 |
| Major cardiovascular diseases, % |  |  |  |  |  |  |
| - No | 7.1 | 6.7 | 7.9 | 0.3 | 0.81 (0.57-1.16) | 0.2 |
| - Yes | 27.9 | 26.2 | 30.5 | 0.4 | 0.98 (0.65-1.48) | 0.9 |
| COPD, % |  |  |  |  |  |  |
| - No | 9.3 | 8.5 | 11.1 | 0.04 | 0.87 (0.65-1.16) | 0.3 |
| - Yes | 28.9 | 27.0 | 33.3 | 0.5 | 0.78 (0.37-1.63) | 0.5 |
| Cancer, % |  |  |  |  |  |  |
| - No | 9.6 | 8.7 | 11.3 | 0.050 | 0.84 (0.63-1.12) | 0.2 |
| - Yes | 20.2 | 18.1 | 26.7 | 0.2 | 0.89 (0.43-1.86) | 0.8 |
| Renal diseases, % |  |  |  |  |  |  |
| - No | 9.3 | 8.6 | 10.9 | 0.07 | 0.93 (0.70-1.24) | 0.6 |
| - Yes | 28.6 | 29.6 | 27.4 | 0.8 | 0.56 (0.27-1.15) | 0.11 |

<sup>a</sup> From March 3 to March 31, 2020.
<sup>b</sup> From April 1 to May 3, 2020.
<sup>c</sup> Chi-squared test for categorical variables, t-test for continuous ones.
<sup>d</sup> Cox proportional hazard model, adjusted for age, gender, hypertension, diabetes, major cardiovascular diseases, COPD, cancer and renal disease. Some models could not be fit due to the scarce number of deaths.

Multivariate Cox analysis did not confirm univariate results: adjusting for age, gender, hypertension, diabetes, CVD, COPD, cancer and renal disease, the hazard ratio (HR) of death of those diagnosed in April, as compared to March, was no more significant: 0.93 (95% Confidence Interval - CI: 0.71-1.21 - Table 2). From March to April, the risk of death did not significantly change in any subset of the sample, with the only exceptions of the females (adjusted HR: 0.65; 95% CI: 0.44-0.95) and diabetics (0.57; 0.34-0.94), whose risk significantly decreased (Table 2). The results of the random-effect logistic regression did not vary: overall, the adjusted odds ratio of death at 28 days was 0.77 (95% CI: 0.55-1.07; p=0.12) for those diagnosed in April, as compared to the subjects detected in March.

## Discussion

In this sample from two Italian provinces, after adjusting for several potential confounders, SARS-CoV-2 case-fatality rate did not significantly decrease from March to April 2020, overall and in any age-class. The mortality trend throughout time was stable across all categories of risk, with the only exceptions of females and diabetics, both showing a significantly lower death rate in April, as compared to March.

Therefore, our data do not support the hypothesis of a decline in SARS-CoV-2 lethality, which has been first suggested by some clinicians [7-11], following a reduction in the number of severe cases requiring hospitalization or ICU support [14]. The above hypothesis, has been recently propelled by the results of a study showing a reduced viral load during the latest phases of the pandemic in the nasopharingeal swabs collected from hospitalized patients in Northern Italy [15]. The available data, however, were based upon a limited number of subjects, and the correlation between viral load and clinical outcome of COVID-19 was almost entirely speculative [15].

In fact, our data are in complete agreement with the available Italian data, showing a substantially stable raw case-fatality rate - approaching 13% over time [16]. It is worth considering, indeed, that none of the potential explanations to support a decline in virus lethality have been demonstrated so far [17]: less virulent virus strains have not been reported [9]; none of the treatments that have been gradually administered to the study participants during the course of the pandemic (including antivirals, low molecular weight heparin and monoclonal antibodies against inflammatory cytokines) has been proven effective to date [18] [19]; finally the healthcare system in the two provinces under study has never been overcrowded during the pandemic [20].

The present data are updating a previous, preliminary analysis [13], significantly expanding the sample and length of follow-up. Yet, they require confirmation from larger datasets across multiple countries, accounting for additional potential confounders (such as body mass index - which we initially did not include among the variables to collect, but later emerged as a significant predictor of death from COVID-19 [21, 22]), during time periods with a stable diagnostic capacity. In fact, in both the selected provinces the number of RT-PCR tests increased substantially from March to April [23], and it cannot be excluded that the proportions of undetected infections or unrecognized COVID-19 deaths were unbalanced over time.

Acknowledging these caveats, our data provide the first evidence of a lack of a significant decrease of SARS-CoV-2 case-fatality rate between March and April, 2020, on a prospective sample, adjusting for several potential predictors of death. We trust that some of the many ongoing trials testing new therapies [24, 25] - starting from those reporting promising results on dexamethasone [26] - will determine a breakthrough in the clinical course of the pandemic.

## Data Availability

All data are available from the corresponding author upon request.

## Acknowledgments

The authors are grateful to Dr. Giorgia Valpiani and Dr. Nicola Napoli for their help in data collection.

## Financial disclosure statement

This work was not funded.

## Competing interests

All authors declare that they have no potential conflict of interest.

## Notes

### Competing Interest Statement

The authors have declared no competing interest.

### Funding Statement

The work was not funded.

### Author Declarations

Ethics Committee of the Emilia Romagna Region.

## References

1. COVID-19 Coronavirus Outbreak [Internet]. Dadax. 2020 [cited March 14, 2020]. Available from: https://www.worldometers.info/coronavirus/.

2. Johns Hopkins University & Medicine. Coronavirus Resource Center - Mortality Analyses 2020 [May 16, 2020]. Available from: https://coronavirus.jhu.edu/data/mortality.

3. Dowd JB, Andriano L, Brazel DM, Rotondi V, Block P, Ding X, et al. Demographic science aids in understanding the spread and fatality rates of COVID-19. Proc Natl Acad Sci U S A. 2020;117(18):9696–8. Epub 2020/04/18. doi:10.1073/pnas.20049111172004911117 [pii]. PubMed PMID: 32300018.

4. Boccia S, Ricciardi W, Ioannidis JPA. What Other Countries Can Learn From Italy During the COVID-19 Pandemic. JAMA internal medicine. 2020. Epub 2020/04/08. doi:10.1001/jamainternmed.2020.1447. PubMed PMID: 32259190.

5. Iosa M, Paolucci S, Morone G. Covid-19: A Dynamic Analysis of Fatality Risk in Italy. Front Med. 2020;7:185. doi:10.3389/fmed.2020.00185.

6. Onder G, Rezza G, Brusaferro S. Case-Fatality Rate and Characteristics of Patients Dying in Relation to COVID-19 in Italy. JAMA. 2020. Epub 2020/03/24. doi:10.1001/jama.2020.46832763667 [pii]. PubMed PMID: 32203977.

7. Caramelo S, Ferreira M, Oliveiros B. Estimation of risk factors for COVID-19 mortality - preliminary results [submitted]. MedRxiv 2020.

8. Lockdown and mutations, the virus is losing its virulence, experts argue [Lockdown e 14 mutazioni, il virus si sta indebolendo]. Il Messaggero. May 7, 2020.

9. Tang X, Wu C, Li X, Song Y, Yao X, Wu X, et al. On the Origin and Continuing Evolution of SARS-CoV-2. National Science Review. 2020;nwaa036. doi:https://doi.org/10.1093/nsr/nwaa036.

10. Di Giambenedetto S, Ciccullo A, Borghetti A, Gambassi G, Landi F, Visconti E, et al. Off-label Use of Tocilizumab in Patients with SARS-CoV-2 Infection. Journal of medical virology. 2020. Epub 2020/04/17. doi:10.1002/jmv.25897. PubMed PMID: 32297987.

11. AIFA. LMWH in adult COVID-19 patients [Eparine a basso peso molecolare nei pazienti adulti con COVID-19]. In: Italian, Medicines, Agency, editors. Rome 2020.

12. Manzoli L, La Vecchia C, Flacco ME, Capasso L, Simonetti V, Boccia S, et al. Multicentric cohort study on the long-term efficacy and safety of electronic cigarettes: study design and methodology. BMC Public Health. 2013;13:883. Epub 2013/09/26. doi:10.1186/1471-2458-13-8831471-2458-13-883 [pii]. PubMed PMID: 24063569; PubMed Central PMCID: PMC3853622.

13. Flacco M, Acuti Martellucci, C. Bravi, F, Parruti, G, Mascitelli, A, Mantovani, L, Manzoli, L. SARS-CoV-2 lethality decreased over 1 time in two Italian Provinces. MedRxiv. 2020. doi:10.1101/2020.05.23.20110882.

14. Italian Institute of Health. COVID-19 integrated surveillance: key national data. Italian Institute of Health, 2020.

15. Clementi N, Ferrarese R, Tonelli M, Amato V, Racca S, Locatelli M, et al. Lower nasopharyngeal viral load during the latest phase of COVID-19 pandemic in a Northern Italy University Hospital. Clin Chem Lab Med. 2020. Epub 2020/07/01. doi:10.1515/cclm-2020-0815/j/cclm.ahead-of-print/cclm-2020-0815/cclm-2020-0815.xml [pii]. PubMed PMID: 32598306.

16. Protezione Civile. COVID-19 Italia - Monitoraggio della situazione 2020 [April 6, 2020]. Available from: http://opendatadpc.maps.arcgis.com/apps/opsdashboard/index.html#/b0c68bce2cce478eaac82fe38d4138b1.

17. MacLean O, Orton RJ, Singer JB, Robertson D. No evidence for distinct types in the evolution of SARS-CoV-2. Virus Evolution. 2020;6(1):veaa034. doi:10.1093/ve/veaa034.

18. NIH.gov. Potential Antiviral Drugs Under Evaluation for the Treatment of COVID-19. NIH, 2020.

19. Wiersinga WJ, Rhodes A, Cheng AC, Peacock SJ, Prescott HC. Pathophysiology, Transmission, Diagnosis, and Treatment of Coronavirus Disease 2019 (COVID-19): A Review. JAMA. 2020. Epub 2020/07/11. doi:10.1001/jama.2020.128392768391 [pii]. PubMed PMID: 32648899.

20. Phua J, Weng L, Ling L, Egi M, Lim CM, Divatia JV, et al. Intensive care management of coronavirus disease 2019 (COVID-19): challenges and recommendations. Lancet Respir Med. 2020;8(5):506–17. Epub 2020/04/10. doi:S2213-2600(20)30161-2 [pii] 10.1016/S2213-2600(20)30161-2. PubMed PMID: 32272080; PubMed Central PMCID: PMC7198848.

21. Rottoli M, Bernante P, Belvedere A, Balsamo F, Garelli S, Giannella M, et al. How important is obesity as a risk factor for respiratory failure, intensive care admission and death in hospitalised COVID-19 patients? Results from a single Italian centre. Eur J Endocrinol. 2020. Epub 2020/07/17. doi:10.1530/EJE-20-0541. PubMed PMID: 32674071.

22. Lighter J, Phillips M, Hochman S, Sterling S, Johnson D, Francois F, et al. Obesity in patients younger than 60 years is a risk factor for Covid-19 hospital admission. Clinical infectious diseases : an official publication of the Infectious Diseases Society of America. 2020. Epub 2020/04/10. doi:10.1093/cid/ciaa415. PubMed PMID: 32271368; PubMed Central PMCID: PMC7184372.

23. Coronavirus, la situazione in Italia [Internet]. Gedi Visual. 2020. Available from: https://lab.gedidigital.it/gedi-visual/2020/coronavirus-i-contagi-in-italia/index.php#tamponi.

24. World Health Organization. Data visualizations and mapping of registered studies for COVID-19 experimental treatments. 2020.

25. Acuti Martellucci C, Flacco, M.E., Cappadona, R., Bravi, F., Mantovani, L., Manzoli, L. SARS-CoV-2 pandemic: An overview. Advances in Biological Regulation. 2020;77. doi:10.1016/j.jbior.2020.100736.

26. Horby P, Lim WS, Emberson JR, Mafham M, Bell JL, Linsell L, et al. Dexamethasone in Hospitalized Patients with Covid-19 - Preliminary Report. The New England journal of medicine. 2020. Epub 2020/07/18. doi:10.1056/NEJMoa2021436. PubMed PMID: 32678530.

